# THE IMPACT OF THE COVID-19 PANDEMIC ON NON-COVID-19 COMMUNITY-ACQUIRED PNEUMONIA, A RETROSPECTIVE COHORT STUDY

**DOI:** 10.1101/2023.05.04.23289541

**Authors:** Terry Lee, Keith R. Walley, John H. Boyd, Kelly A. Cawcutt, Andre C. Kalil, James A. Russell

**Affiliations:** Centre for Health Evaluation and Outcome Science (CHEOS), St. Paul’s Hospital and University of British Columbia, 1081 Burrard Street, Vancouver, BC, Canada V6Z 1Y6; Centre for Heart Lung Innovation, St. Paul’s Hospital, University of British Columbia, Vancouver, BC, Canada; Division of Critical Care Medicine, St. Paul’s Hospital, University of British Columbia, Vancouver, BC, Canada; University of Nebraska Medical Center, Omaha, Nebraska, USA

**Keywords:** community-acquired pneumonia, CAP, coronavirus, COVID-19, SARS-CoV-2, pneumococcus, influenza

## Abstract

**BACKGROUND:** The COVID-19 pandemic could impact frequency and mortality of non-COVID-19 community-acquired pneumonia (CAP). Changes in frequency, patient mix, treatment, and organ dysfunction could cascade together to increase mortality of CAP during compared to pre-COVID-19.

**METHODS:** Hospitalized CAP patients at St. Paul’s Hospital, Vancouver, Canada pre- (fiscal years 2018/2019 and 2019/2020) and during COVID-19 pandemic (2020/2021 and 2021/2022) were evaluated.

**RESULTS:** In 5219 CAP patients, there was no significant difference pre-versus during pandemic in mean age, gender and Charlson co-morbidity score. However, hospital mortality increased significantly from pre-versus during COVID-19 (7.5% versus 12.1% respectively, [95% CI for difference: 3.0-6.3%], p<0.001), a 61% relative increase, coincident with increases in ICU admission (18.3% versus 25.5% respectively, [95% CI for difference: 5.0-9.5%] p<0.001, 39% relative increase) and ventilation (12.7% versus 17.5%, respectively, [95% CI for difference: 2.8-6.7%] p<0.001, 38% relative increase). Results remained the same after regression adjustment for confounders. CAP hospital admissions decreased 27% from pre- (n=1349 and 1433, 2018/2019 and 2019/2020 respectively) versus the first COVID-19 pandemic year (n=1047 in 2020/2021) then rose to pre-pandemic number (n=1390 in 2021/2022). During pre-pandemic years, CAP admissions peaked in winter; during COVID-19, the CAP admissions peaked every six months.

**CONCLUSIONS AND RELEVANCE:** The COVID-19 pandemic was associated with increases in hospital mortality, ICU admission and invasive mechanical ventilation rates of non-COVID-19 CAP and a transient, one year frequency decrease. There was no winter seasonality of CAP during the COVID-19 pandemic era. Future pandemic planning for CAP hospital care is needed.

**What is already known on this topic:** The COVID-19 pandemic could impact frequency and mortality of non-COVID-19 community-acquired pneumonia (CAP). No prior study has examined this hypothesis.

**What this study adds:** The COVID-19 pandemic was associated with increases in hospital mortality, ICU admission and invasive mechanical ventilation rates of non-COVID-19 CAP and a transient, one year frequency decrease. There was no winter seasonality of CAP during the COVID-19 pandemic era.

**How this study might affect research, practice or policy:** Future pandemic planning for CAP hospital care is needed.

## INTRODUCTION

The COVID-19 pandemic could have impacted non-COVID-19 community-acquired pneumonia (CAP herein)(1). CAP is the commonest lethal infection affecting > 1% of the population causing 17-23 deaths per 100,000 population in Canada in 2016-2020(2). In the USA, deaths due to “influenza and pneumonia” increased from 49,783 pre-COVID-19 (2019) to 53,495 during COVID-19 (2020)(3).

During the COVID-19 pandemic, several factors could potentially decrease CAP frequency and mortality while other opposing factors could increase CAP frequency and mortality.

The frequency of CAP can be *decreased* by out-patient interventions that can decrease numbers of patients requiring hospitalization including vaccines (e.g. influenza(4), pneumococcal), by social measures (e.g., hand-washing and social distancing(5)), by optimal treatment of underlying co-morbidities(6), and initiation of out-patient antimicrobials. COVID-19 could also *decrease* frequency of CAP because of decreased frequency of influenza, a precursor of CAP, in Canada(7). Pneumococcal CAP decreased by 30% in the UK during COVID-19(8).

Hospital mortality of CAP can be *decreased* by optimizing hospital management, including early diagnosis and antimicrobial treatment(6), oxygen supplementation, and in the critically ill, mechanical ventilation, vasopressors, corticosteroids(9, 10) for vasopressor-resistant septic shock, and renal support for acute kidney injury(6, 11). Very recently, hydrocortisone was shown to decrease mortality of critically ill CAP patients(12).

The COVID-19 pandemic could *increase* CAP frequency and/or mortality because of inadequate treatment of comorbidities that increase risk of CAP(13-16), increased alcohol(17) and drug(18) use, decreased use of pneumococcal vaccine(19), avoidance of clinic, ED(20, 21) (down by 50% at COVID-19 peak(22)) and hospital admissions because of fear of contracting COVID-19, increased use of virtual medicine (e.g. from 1% to 29% in Ontario(23, 24)), delays in hospital admission and treatment due to hospital overcrowding, and inadequate adherence to CAP treatment guidelines(6) because of shortages of staff and caregiver fatigue and shortages.

Our hypothesis was that changes in frequency, patient mix, treatment, and organ dysfunction cascaded together to increase mortality of CAP during COVID-19 compared to pre-COVID-19. We sought to compare CAP frequency, baseline characteristics, comorbidities, treatment including ICU admission and ventilation use and hospital mortality rates two years immediately pre-COVID-19 pandemic versus the first two years of the pandemic. Using adjusted analyses, we aimed to determine whether any changes in hospital mortality of CAP were explained by the differences in baseline characteristics and treatment.

## METHODS

### Study design and participants

This study was a single hospital in-patient electronic medical record Cerner database retrospective cohort study, reported in accordance with the STROBE checklist(25). This study was approved by the Providence Health Care and University of British Columbia (UBC) Human Research Committee as low risk and no consent was required.

We reviewed the St. Paul’s Hospital (Vancouver, Canada) Cerner electronic medical records of CAP admissions during four fiscal years between April 1, 2018 and March 31, 2022. Non-COVID-19 CAP was defined by ICD10 codes for viral, bacterial and aspiration CAP (**Supplement Table 1**). We excluded Emergency Department only admission, CAP readmissions, and patients who were COVID-19 SARS-CoV-2 positive at admission.

At admission we recorded, age, sex, Charlson comorbidity score, mean, systolic and diastolic arterial pressures, heart and respiratory rates, arterial pulse oximetry and temperature. Vital signs were only available on Cerner electronic medical records for patients admitted after November 15, 2019.

The primary outcome was hospital mortality; secondary outcomes were ICU admission, use of mechanical ventilation, time to death and duration of hospital length of stay. We also examined the seasonality of frequency, hospital mortality, ICU admission and use of mechanical ventilation by fiscal year quarters

### Statistical Analyses

Patients’ age, sex, co-morbidities (Charlson score), admission day vital signs (available for patients admitted after November 15, 2019), dates of admission and discharge or death were compared for the two pre-COVID-19 pandemic years to the first two COVID-19 pandemic years (2018/2019 and 2019/2020 versus 2020/2021 and 2021/2022). Age, sex, co-morbidities, and vital signs were compared pre-versus during pandemic by ANOVA, Kruskal–Wallis test or Chi-square test as appropriate, as were for binary and continuous outcomes. Kaplan-Meier curves and log rank test were used to compare time to death. Patients discharged alive were censored at the time of hospital discharge. Analysis adjusted for age, sex and Charlson score was performed by logistic or Cox regression as appropriate. Results were expressed as odds ratio (OR) and hazard ratio (HR) with 95% confidence interval.

## RESULTS

In 5219 CAP admissions, patients in pre-versus during pandemic had higher admission mean arterial pressure (median (IQR): 90 (80, 103) versus 88 (78, 101), mmHg [95% CI for difference: 0.2-3.8] p=0.015), higher systolic blood pressure (127 (111, 144) versus 124 (110, 141) mmHg, [95% CI for difference: 0.5-5.5] p=0.047), higher arterial oxygen saturation (96 (94, 98) versus 95 (93, 97), [95% CI for difference: 0.7-1.3], p=0.009), and faster heart rate (98 (83, 115) versus 95 (80, 110) beast/minute, [95% CI for difference: 0.1-5.9] p=0.002)(**Table 1**), but no differences in comorbidities, gender, age or admission respiratory rate, diastolic blood pressure, or temperature pre-versus during pandemic (**Table 1**).

**Table 1.**
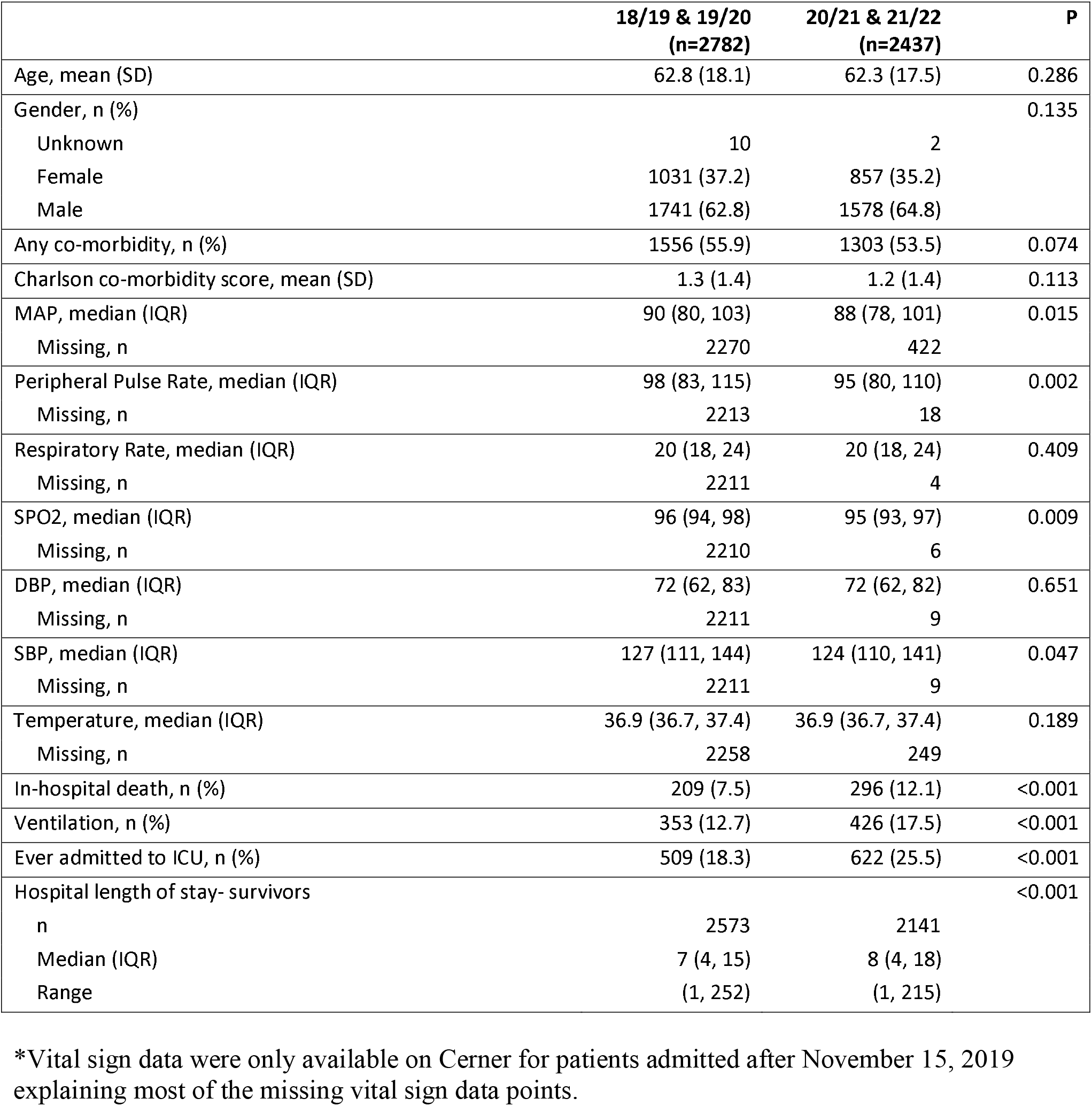
Baseline characteristics and unadjusted outcomes of non-COVID-19 CAP for the two years pre-and the first two years during the COVID-19 pandemic*.

|  | **18/19 & 19/20**<br>**(n=2782)** | **20/21 & 21/22**<br>**(n=2437)** | **P** |
| --- | --- | --- | --- |
| Age, mean (SD) | 62.8 (18.1) | 62.3 (17.5) | 0.286 |
| Gender, n (%) |  |  | 0.135 |
| Unknown | 10 | 2 |  |
| Female | 1031 (37.2) | 857 (35.2) |  |
| Male | 1741 (62.8) | 1578 (64.8) |  |
| Any co-morbidity, n (%) | 1556 (55.9) | 1303 (53.5) | 0.074 |
| Charlson co-morbidity score, mean (SD) | 1.3 (1.4) | 1.2 (1.4) | 0.113 |
| MAP, median (IQR) | 90 (80, 103) | 88 (78, 101) | 0.015 |
| Missing, n | 2270 | 422 |  |
| Peripheral Pulse Rate, median (IQR) | 98 (83, 115) | 95 (80, 110) | 0.002 |
| Missing, n | 2213 | 18 |  |
| Respiratory Rate, median (IQR) | 20 (18, 24) | 20 (18, 24) | 0.409 |
| Missing, n | 2211 | 4 |  |
| SPO2, median (IQR) | 96 (94, 98) | 95 (93, 97) | 0.009 |
| Missing, n | 2210 | 6 |  |
| DBP, median (IQR) | 72 (62, 83) | 72 (62, 82) | 0.651 |
| Missing, n | 2211 | 9 |  |
| SBP, median (IQR) | 127 (111, 144) | 124 (110, 141) | 0.047 |
| Missing, n | 2211 | 9 |  |
| Temperature, median (IQR) | 36.9 (36.7, 37.4) | 36.9 (36.7, 37.4) | 0.189 |
| Missing, n | 2258 | 249 |  |
| In-hospital death, n (%) | 209 (7.5) | 296 (12.1) | <0.001 |
| Ventilation, n (%) | 353 (12.7) | 426 (17.5) | <0.001 |
| Ever admitted to ICU, n (%) | 509 (18.3) | 622 (25.5) | <0.001 |
| Hospital length of stay- survivors |  |  | <0.001 |
| n | 2573 | 2141 |  |
| Median (IQR) | 7 (4, 15) | 8 (4, 18) |  |
| Range | (1, 252) | (1, 215) |  |

\*Vital sign data were only available on Cerner for patients admitted after November 15, 2019 explaining most of the missing vital sign data points.

CAP hospital admission frequency decreased 27% pre-versus during pandemic (1047 in 2020/2021 versus 1349 and 1433 in 2018/2019 and 2019/2020 respectively) then returned to pre-pandemic levels (1390 in 2021/2022) (**Figure 1**).

**Figure 1.**
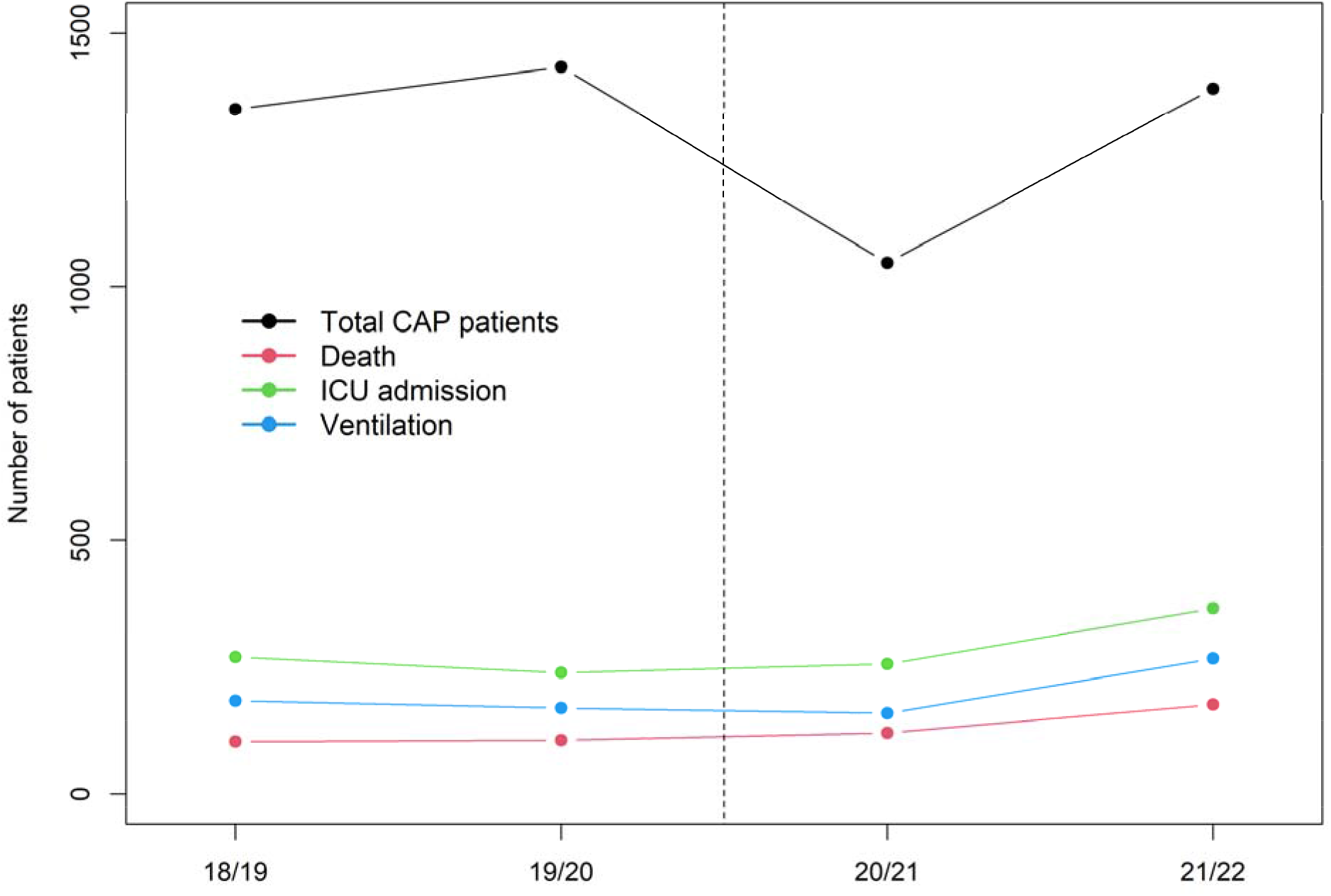
CAP frequency pre-versus during the pandemic. Numbers of CAP admissions are shown relative to fiscal years. The dashed line indicates when we defined pandemic onset, comparing two years pre-pandemic to the first two years of the pandemic.

The first two years of the COVID-19 pandemic was associated with increased absolute numbers of deaths compared to the two years pre-COVID-19 pandemic, increasing from 209 in the two pre-COVID-19 pandemic years to 296 in the first two years of the pandemic. The absolute number of CAP deaths per year increased in the second COVID-19 pandemic year fiscal year; from 103, 106, and 120 in fiscal years 2018/2019, 2019/2020 and 2020/2021 to 176 in fiscal year 2021/2022 (**Figure 1)**

Hospital mortality increased significantly pre-versus during pandemic (7.5% versus 12.1% respectively, [95% CI for difference: 3.0-6.3%] p< 0.001; 61% relative increase), coincident with significant increases in ICU admission (18.3% versus 25.5% respectively, [95% CI for difference: 5.0-9.5%] p<0.001; 39% relative increase) and ventilation (12.7% versus 17.5%, respectively, [95% CI for difference:2.8-6.7%] p<0.001; 38% relative increase) (**Table 1)**.

Survival times of pre-versus during the COVID-19 pandemic decreased significantly (**Figure 2B**; HR: 1.39 (95% CI: 1.17, 1.66), p<0.001). Notably, the survival curves of pre-versus during the pandemic separated at about 12 days in the pre-versus during and then continued to separate indicating lower mortality after 12 days in the pre-versus during pandemic CAP patients.

**Figure 2.**
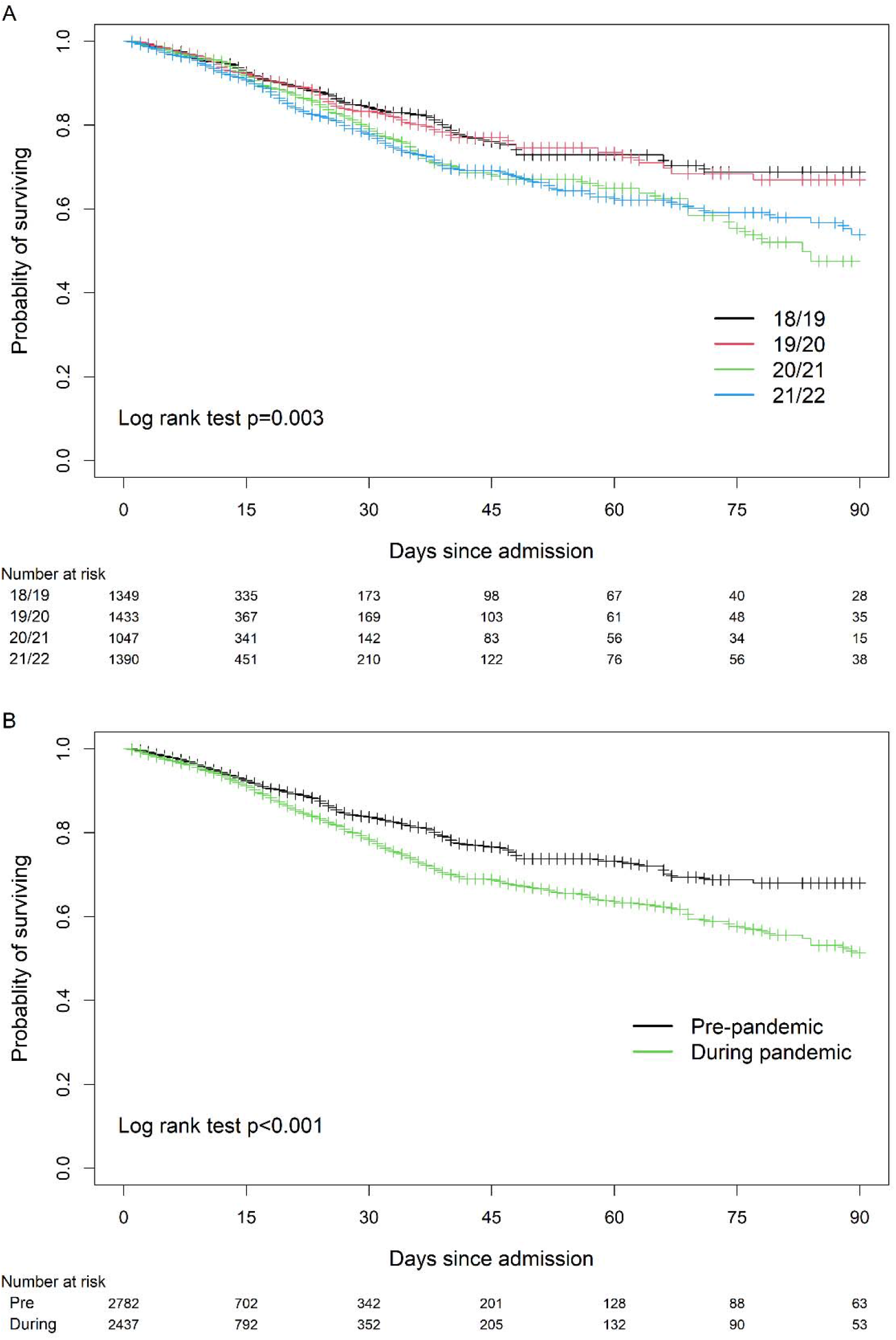
A) CAP hospital mortality pre-versus during the pandemic by fiscal year and according to the 2 years pre-versus 2 years during the pandemic B) according to the pooled two fiscal years pre-COVID-19 pandemic (2018/2019 and 2019/2020) versus pooled two fiscal years during the COVID-19 pandemic (2020/2021 and 21021/2022).

The CAP survivors’ length of stay also changed during the COVID-19 pandemic – during the COVID-19 pandemic CAP patients had longer hospital length of stay than pre-COVID-19 CAP patients (**Table 1**).

Hospital mortality, ICU admission and mechanical ventilation rates remained significantly different for pre-versus during pandemic in adjusted regression analyses (**Table 2**).

**Table 2.** Outcomes (time to death, in-hospital death, use of mechanical ventilation and ever admitted to ICU during hospital admission) adjusted for age, sex, and co-morbidities.

|  | Unadjusted analysis |  | Adjusted analysis |  |
| --- | --- | --- | --- | --- |
|  | Hazard ratio or odds ratio (95% CI) | P | Hazard ratio or odds ratio (95% CI) | P |
| Time to death |  |  |  |  |
| 19/20 vs. 18/19 | 1.00 (0.76, 1.31) | 0.999 | 1.06 (0.81, 1.40) | 0.659 |
| 20/21 vs. 18/19 | 1.35 (1.04, 1.76) | 0.024 | 1.39 (1.07, 1.81) | 0.015 |
| 21/22 vs. 18/19 | 1.42 (1.12, 1.81) | 0.005 | 1.52 (1.19, 1.94) | <0.001 |
| During vs. pre-pandemic | 1.39 (1.17, 1.66) | <0.001 | 1.42 (1.19, 1.69) | <0.001 |
| In-hospital death |  |  |  |  |
| 19/20 vs. 18/19 | 0.97 (0.73, 1.28) | 0.812 | 1.01 (0.75, 1.35) | 0.959 |
| 20/21 vs. 18/19 | 1.57 (1.19, 2.07) | 0.001 | 1.59 (1.20, 2.12) | 0.001 |
| 21/22 vs. 18/19 | 1.75 (1.36, 2.26) | <0.001 | 1.94 (1.49, 2.53) | <0.001 |
| During vs. pre-pandemic | 1.70 (1.41, 2.05) | <0.001 | 1.78 (1.47, 2.15) | <0.001 |
| Ventilation |  |  |  |  |
| 19/20 vs. 18/19 | 0.85 (0.68, 1.06) | 0.144 | 0.83 (0.66, 1.04) | 0.104 |
| 20/21 vs. 18/19 | 1.13 (0.90, 1.43) | 0.284 | 1.12 (0.89, 1.42) | 0.318 |
| 21/22 vs. 18/19 | 1.51 (1.23, 1.85) | <0.001 | 1.47 (1.20, 1.81) | <0.001 |
| During vs. pre-pandemic | 1.46 (1.25, 1.70) | <0.001 | 1.45 (1.24, 1.69) | <0.001 |
| Ever admitted to ICU |  |  |  |  |
| 19/20 vs. 18/19 | 0.80 (0.66, 0.97) | 0.023 | 0.78 (0.64, 0.95) | 0.013 |
| 20/21 vs. 18/19 | 1.29 (1.07, 1.57) | 0.009 | 1.29 (1.06, 1.56) | 0.012 |
| 21/22 vs. 18/19 | 1.43 (1.19, 1.71) | <0.001 | 1.39 (1.16, 1.67) | <0.001 |
| During vs. pre-pandemic | 1.53 (1.34, 1.75) | <0.001 | 1.52 (1.33, 1.74) | <0.001 |

The seasonality of frequency, hospital mortality, ICU admission and mechanical ventilation use was evaluated by examining each of these variables according to fiscal year quarter (**Figure 3**). There was seasonality of CAP frequency that differed in the pre-COVID-19 pandemic versus during COVID-19 pandemic eras. During the pre-COVID-19 pandemic years, CAP admissions peaked annually in winter quarters whereas during COVID-19 years, the CAP frequency peaked about every six months (**Figure 3**). There was no seasonality of use of ventilation or hospital mortality rates expressed as percentages. Interestingly, the ICU admission rates (percentages) peaked in the second quarters of each of the four years of study, in the pre- (Q2 2018 and Q2 2019) and during COVID-19 pandemic years (Q2 2020 and Q2 2021, **Figure 3B**).

**Figure 3.**
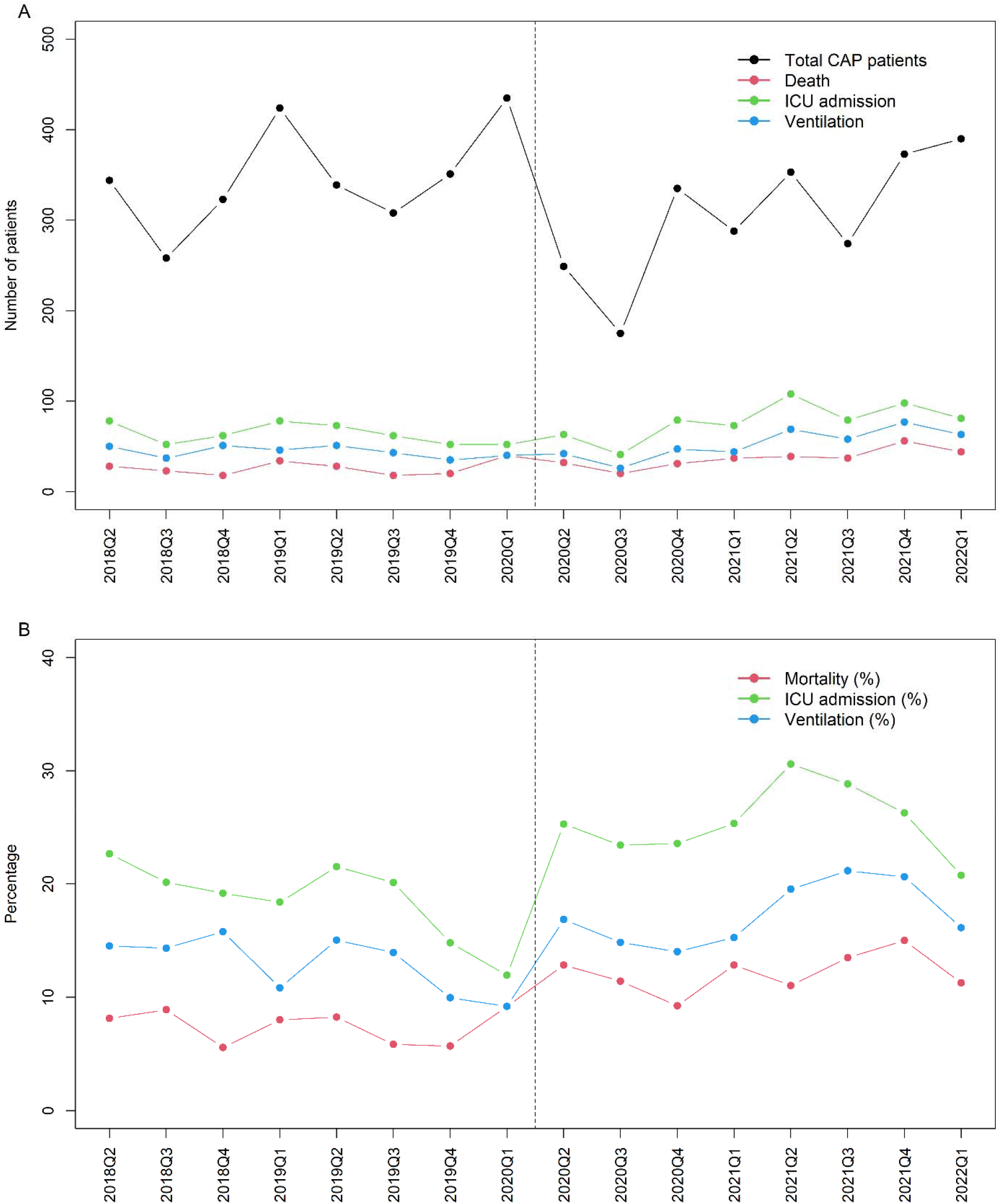
Seasonality of CAP frequency, hospital mortality, ICU admissions and use of invasive mechanical ventilation by fiscal year quarter.

## DISCUSSION

In 5219 CAP admissions, there was a significant 61% relative increase of hospital mortality during versus pre-pandemic and 40% relative increases of ICU admission and invasive ventilation rates. Pre-pandemic CAP winter seasonality preceded a transient 27% decrease in frequency in the first pandemic year. The COVID-19 pandemic appeared to change the seasonality of CAP; CAP frequency peaked annually in the winter months pre-pandemic, but then peaked every six months during the first two pandemic years.

There were no differences in age, sex, or co-morbidities to explain increases in CAP outcomes; adjusted analyses yielded the same outcome findings. Small statistically significant differences in admission mean and systolic arterial pressure and heart rate were not clinically relevant, reflecting the large sample size.

There are no publications formally assessing the impact of the COVID-19 pandemic on hospital mortality of CAP. There is some information about overall CAP mortality before versus during COVID-19. COVID-19 mortality was higher than influenza mortality in studies of influenza pre-pandemic(28, 29) and deaths due to “influenza and pneumonia” increased in the USA from 49,783 in 2020 compared to 53,495 in 2019 (3).

Although we found that the COVID-19 pandemic had specific effects on CAP that required hospital admission, to date, there are few studies of the effects of the COVID-19 era on overall CAP frequency and incidence. Several other studies suggest that overall out-patient and hospitalized CAP may have decreased during the pandemic in Japan(26), Canada (significant decreases in incidence of influenza and respiratory syncytial virus during versus before COVID-19(7)), the UK(8) and other countries(6, 26, 27).

We found that the number of hospital admissions for CAP decreased in the first year of the pandemic compared to pre-pandemic, but the reasons are not clear. Factors that could decrease CAP frequency include CAP vaccines such as pneumococcal and influenzas vaccines. Unfortunately, we did not have such data. In Japan, pneumococcal vaccine use decreased yet invasive pneumococcal disease in children was lower during versus COVID-19 due to containment measures (19).

Pandemic social interventions (e.g. mandatory masking, lockdowns and travel restrictions; increased use of virtual medicine; inadequate treatment of chronic conditions) could have decreased the number of CAP admissions. Decreased seasonal respiratory infection during versus before COVID-19 was attributed to social interventions in Canada (7), in a multi-country study (sustained decrease in invasive *S. pneumoniae, H. influenzae*, and *N. meningitidis*(30)), in Maryland USA (mask mandates were associated with decreased Emergency Department visits for viral disease, asthma and COPD exacerbations, common risk factors for CAP(21)) and in Scandinavia (decreased frequency of influenza in the first pandemic year(5)).

It would appear that factors that could have increased hospital admissions for CAP during the pandemic might not have been dominant enough to offset the above factors that could have decreased CAP admissions during the first pandemic year. For example, alcohol and drug use are risk factors for CAP that could have altered pandemic versus pre-pandemic frequency of CAP, but we do not have such information in our cohort. Alcohol and drug use increased in the US(31, 32) but varied by country(33) during versus before COVID-19 pandemic catalyzing CAP(34) risk.

We found that hospital mortality increased by 61% relatively in the first two pandemic versus the two immediate pre-pandemic years. COVID-19 could have increased mortality of hospitalized CAP because of inadequate comorbidity management (e.g. heart failure, COPD, chronic kidney disease and diabetes(13-15)), decreased health care visits for fear of contracting COVID-19 (4), increased use of virtual medicine, decreased adherence to treatment guidelines(6, 35), decreased hospital and ICU bed availability because of over-crowding.

We found that there was no difference in frequency of co-morbidities pre-versus during the pandemic and in adjusted analyses, the primary outcome, hospital mortality rate of CAP, remained significantly higher during than pre-pandemic. Comorbidities that increase risk of CAP include hypertension, asthma, heart failure, COPD, chronic kidney disease and diabetes(13-15). Effective chronic condition management mitigates the risk of getting CAP and hospital admission (14).

Virtual medicine use could have altered CAP frequency in our study. In British Columba, Canada virtual medicine use increased nearly 400% during the pandemic (28 to 106 visits/physician April 2019 to March 2021 (36). Virtual medicine dramatically increased in Ontario, Canada in the early pandemic era virtual care increased from 1.6% of out-patient visits (second quarter 2019) to 70.6% (second quarter of 2020)(23).

We were not able to evaluate compliance with CAP treatment guidelines(6, 35) in this hospital database retrospective cohort study. We were also unable to obtain bed availability pre-versus during pandemic. During COVID-19 surges, over-crowding of hospitals(37-39) limited accessibility for patients with CAP. Furthermore, many countries identified lack of ICU beds and ventilators(39) prior to COVID-19 (40). Patients could have feared visiting medical facilities for fear of contracting COVID-19 (20, 21).

We believe this is the first report that the COVID-19 pandemic had clinically relevant effects on hospitalized CAP patients’ mortality, ICU admission and invasive ventilation and frequency. Understanding the effects of the COVID-19 pandemic on CAP could be used for future pandemic preparedness because most pandemics are respiratory infections that cause CAP. There are well-known seasonal variations in CAP due to influenza and common bacterial causes of CAP, both non-COVID-19 CAP and pandemic causes of CAP are altered by prevention (treatment of underlying conditions that are risk factors and vaccines) and non-specific supportive treatment (oxygen, vital organ support and ICU admission) and specific antimicrobials and host response interventions (such as anti-inflammatory agents).

Strengths of our study are the use of a comprehensive Cerner hospital database to identify and include CAP patients, and to evaluate frequency and outcomes consistently in the two immediate pre-pandemic years versus the first two pandemic years, and that our primary outcome was robust to adjustment for age, sex and comorbidities.

Limitations of our study include that this is a retrospective single centre cohort study, that we used ICD10 codes to identify CAP patients, and that we did not have data to evaluate the effects of societal interventions such as lockdowns and travel restrictions, data on vaccine use or telemedicine use, CAP treatment guideline compliance, or hospital bed availability.

In conclusion, hospital mortality, ICU admission and invasive mechanical ventilation rates of non-COVID-19 CAP increased significantly during the COVID-19 pandemic. A transient one year decrease of CAP frequency was followed by six-monthly rather than winter CAP frequency peaks. This can inform future pandemic planning for hospitalized CAP.

## Data Availability

All data produced in the present work are contained in the manuscript

## Figure Legends

**Figure 1.** The absolute numbers of CAP cases in St. Paul’s Hospital, Vancouver, BC, Canada are shown according to the fiscal years pre-COVID-19 pandemic (2018/2019 and 2019/2020) and during the COVID-19 pandemic (2020/2021 and 21021/2022).

**Figure 2:** The hospital mortality rates of CAP cases in St. Paul’s Hospital, Vancouver, BC, Canada are shown in 2A according to each of the fiscal years 2018/2019, 2019/2020, 2020/2021 and 21021/2022 and in 2B according to the pooled two fiscal years pre-COVID-19 pandemic (2018/2019 and 2019/2020) versus pooled two fiscal years during the COVID-19 pandemic (2020/2021 and 21021/2022).

**Figure 3.** Seasonality of CAP frequency, ICU admission, use of mechanical ventilation and hospital mortality of CAP cases in St. Paul’s Hospital, Vancouver, BC, Canada in the pre- and during COVID-19 pandemic years. The absolute numbers of cases, ICU admissions, numbers needing ventilation and numbers of deaths are shown in Figure 3A according to quarter. The ICU admission, ventilation and mortality rates expressed as percentages are shown in Figure 3B according to quarter.

## Competing Interests Statement

Drs. Lee, Boyd and Kalil declare that they have no conflict of interest.

Dr Walley has received Foundation Grant from the Canadian Institutes for Health Research, held by UBC. He is the Chair of a DSMB for Northern Therapeutics, unpaid service.

Dr Cawcutt received payment from Becton, Dickinson and Company for advisory meeting participation and speaking related to sepsis from October 2022.

Dr. Russell reports patents owned by the University of British Columbia (UBC) that are related to (1) the use of PCSK9 inhibitor(s) in sepsis, (2) the use of vasopressin in septic shock and (3) a patent owned by Ferring for use of selepressin in septic shock. Dr. Russell is an inventor on these patents. Dr. Russell was a founder, Director and shareholder in Cyon Therapeutics Inc. (now closed) and is a shareholder in Molecular You Corp. Dr. Russell is Senior Research Advisor of the British Columbia, Canada Post COVID – Interdisciplinary Clinical Care Network (PC-ICCN).

Dr. Russell is no longer actively consulting for any industry.

Dr. Russell reports receiving consulting fees in the last 3 years from:

1. Dr. Russell was a funded member of the Data and Safety Monitoring Board (DSMB) of an NIH-sponsored trial of plasma in COVID-19 (PASS-IT-ON) (2020-2021).
2. PAR Pharma (sells prepared bags of vasopressin).

Dr. Russell has received grants for COVID-19 and for pneumonia research: 4 from the Canadian Institutes of Health Research (CIHR) and 3 from the St. Paul’s Foundation (SPF).

Dr. Russell was a non-funded Science Advisor and member, Government of Canada COVID-19 Therapeutics Task Force (June 2020 – 2021).

**Supplement Table 1.** ICD-10 codes used to identify CAP.

| **DxId** | **DxCode** | **DxDesc** |
| --- | --- | --- |
| 105 | A403 | Sepsis due to Streptococcus pneumoniae |
| 362 | B953 | Streptococcus pneumoniae as the cause of diseases classified to other chapters |
| 369 | B960 | Mycoplasma pneumoniae [M. pneumoniae] as the cause of diseases classified to other chapters |
| 370 | B961 | Klebsiella pneumoniae [K. pneumoniae] as the cause of diseases classified to other chapters |
| 3051 | J100 | Influenza with pneumonia, other influenza virus identified |
| 3054 | J110 | Influenza with pneumonia, virus not identified |
| 3057 | J120 | Adenoviral pneumonia |
| 3058 | J121 | Respiratory syncytial virus pneumonia |
| 3059 | J122 | Parainfluenza virus pneumonia |
| 3060 | J128 | Other viral pneumonia |
| 3061 | J129 | Viral pneumonia, unspecified |
| 3062 | J13 | Pneumonia due to Streptococcus pneumoniae |
| 3063 | J14 | Pneumonia due to Haemophilus influenzae |
| 3064 | J150 | Pneumonia due to Klebsiella pneumoniae |
| 3065 | J151 | Pneumonia due to Pseudomonas |
| 3066 | J152 | Pneumonia due to Staphylococcus |
| 3067 | J153 | Pneumonia due to Streptococcus, group B |
| 3068 | J154 | Pneumonia due to other streptococci |
| 3069 | J155 | Pneumonia due to Escherichia coli |
| 3070 | J156 | Pneumonia due to other aerobic Gram-negative bacteria |
| 3071 | J157 | Pneumonia due to Mycoplasma pneumoniae |
| 3072 | J158 | Other bacterial pneumonia |
| 3073 | J159 | Bacterial pneumonia, unspecified |
| 3074 | J168 | Pneumonia due to other specified infectious organisms |
| 3075 | J170 | Pneumonia in bacterial diseases classified elsewhere |
| 3076 | J171 | Pneumonia in viral diseases classified elsewhere |
| 3077 | J172 | Pneumonia in mycoses |
| 3078 | J173 | Pneumonia in parasitic diseases |
| 3079 | J178 | Pneumonia in other diseases classified elsewhere |
| 3080 | J180 | Bronchopneumonia, unspecified |
| 3081 | J181 | Lobar pneumonia, unspecified |
| 3082 | J182 | Hypostatic pneumonia, unspecified |
| 3083 | J188 | Other pneumonia, organism unspecified |
| 3084 | J189 | Pneumonia, unspecified |
| 3085 | J200 | Acute bronchitis due to Mycoplasma pneumoniae |
| 3192 | J851 | Abscess of lung with pneumonia |
| 3193 | J852 | Abscess of lung without pneumonia |
| 29991 | J123 | Human metapneumovirus pneumonia |
| 30006 | J160 | Chlamydial pneumonia |
| 39540 | U002 | Infection with penicillin-resistant *Streptococcus pneumoniae* |

